# Systematic review of spontaneous reports of myocarditis and pericarditis in transplant recipients and immunocompromised patients following COVID-19 mRNA vaccination

**DOI:** 10.1101/2021.12.20.21268102

**Authors:** Samantha Lane, Alison Yeomans, Saad Shakir

**Affiliations:** Drug Safety Research Unit, Southampton, UK; School of Pharmacy & Biomedical Sciences, University of Portsmouth, UK

## Abstract

**Objectives:** To determine whether spontaneous reporting rates of myocarditis and pericarditis differed in immunocompromised patients compared to the whole population overall, and in terms of demographics, vaccine dose, and time-to-onset.

**Design:** Systematic review of spontaneously reported data from the European Union/European Economic Area (EU/EEA), the United States (US) and the United Kingdom (UK).

**Data Sources:** EudraVigilance (EU/EEA), Vaccine Adverse Event Reporting System (VAERS; US) and the Medicines and Healthcare products Regulatory Agency (MRHA, UK) spontaneous reporting databases were searched from date of vaccine launch to 01 December 2021.

**Eligibility criteria:** Publicly available spontaneous reporting data for “Myocarditis” and “Pericarditis” from EU/EEA and US following COVID-19 mRNA vaccines. Reports with comorbidities or concurrent medication indicative of transplantation, HIV infection, or cancer (“immunocompromised” population) were compared with each overall database population.

**Data extraction and synthesis:** Two researchers extracted data. Spontaneously reported events of myocarditis and pericarditis were presented for immunocompromised populations for each data source, stratified by age, sex, dose, and time-to-onset (where available). Seriousness of each event was determined according to the ICH E2A definition. Proportional Reporting Ratio (PRR) was calculated.

**Results:** There were 178 reports of myocarditis and pericarditis amongst immunocompromised individuals overall. Seriousness was comparable between the immunocompromised and overall populations in both databases. No trends in age or sex were observed amongst immunocompromised individuals. Most reports followed a second vaccine dose and occurred within 14 days. The frequency of reporting was similar to the wider population (PRR=1.36 [95% CI= 0.89-1.82] for VAERS population).

**Conclusions:** Myocarditis and pericarditis following COVID-19 vaccination are very rare, and benefits of COVID-19 vaccination continue to outweigh any perceived risks. Reporting rates of myocarditis and pericarditis were similar in immunocompromised individuals, however defining characteristics differed compared to the whole population; therefore, continued monitoring of adverse events following vaccination remains vital to understand differences between population subgroups.

**Strengths and Limitations of the Study:**

- This is the first study to bring together spontaneous reporting data from three regions (Europe, the United States, and the United Kingdom) comparing immunocompromised and immunocompetent populations adverse reactions following COVID-19 mRNA vaccination
- Spontaneously reported adverse drug reaction data is known to be subject to underreporting and missing information, including information on comorbidities and concomitant medications.
- Further biases that may have influenced results, include differences in vaccination strategies between the three regions examined, differences in data collected via spontaneous reporting systems, and the fact that serious events are more likely to be reported.
- It is not possible to estimate incidence rates using spontaneous reporting data due to a lack of precise denominator data, i.e. the number of people who received the vaccine in the corresponding period.

## Introduction

The COVID-19 mRNA vaccines developed by Pfizer/BioNTech and Moderna were the first mRNA vaccines approved for use and have been successfully implemented in the fight against the COVID-19 pandemic (1-3). They are successful both in terms of efficacy of the vaccines to elicit a protective immune response as well as the speed of development; this methodology for vaccination likely enables rapid modifications to fight any potential variants that may evade the current vaccinations in the future. This success does not come without concerns though, and careful monitoring in real-world studies are essential to ensure these new mRNA vaccinations are safe across the whole population. It is also important to determine whether multiple dosing has any adverse effects, especially if modified mRNA vaccinations may be used in the future to fight new COVID-19 variants or other viral infections.

Spontaneous reporting of adverse events enables a snapshot of real-world responses to be assessed, with the caveat that there is often under-reporting and missing data within these reports (4, 5). Reported adverse events have been reported across the world in response to COVID-19 mRNA vaccinations and several have been recognised within the product information guidance, including myocarditis, pericarditis, and myopericarditis (6, 7). Myocarditis and pericarditis are caused by inflammation of the myocardium pericardium leading to chest pain, shortness of breath, palpitations, cardiac failure, or abnormal heart rhythms (8, 9). In Europe, myocarditis and pericarditis have been reported at an excess of 26 to 57 events per million within one week of COVID-19 mRNA vaccination, while in the US it has been estimated that these events are reported at a rate of between one and 40.6 cases per million second doses of COVID-19 mRNA vaccines administered (10, 11). Within these populations, reporting rates were dependent on sex and age, with males under 30 years of age reporting myocarditis and pericarditis following COVID-19 mRNA vaccines at a higher rate than females under 30 and people aged 30 years and over (10, 11). The incidence of myocarditis and pericarditis following COVID-19 mRNA vaccination is likely to be higher considering the well-known under-reporting of spontaneous adverse reactions.

Due to potential differences in the immune response to vaccination between immunocompromised and ‘healthy’ individuals, we hypothesised that immunocompromised individuals might experience myocarditis or pericarditis at a higher frequency, that characteristics (age and sex) of immunocompromised patients reporting these events may differ, that clinical course may be more severe, and that duration of symptoms may be longer compared with immune competent individuals. We therefore aimed to determine the number of reports of myocarditis or pericarditis submitted by immunocompromised patient subgroups and whether these reports follow a similar reporting trend, in terms of age, sex, time-to-onset, and vaccine dose, compared with the total population following mRNA vaccination.

## Materials and Methods

Systematic searches of spontaneous reporting outputs of the European Union/European Economic Area (EU/EEA; EudraVigilance) and the United States (US; Vaccine Adverse Event Reporting System [VAERS] via CDC Wonder tool) were conducted to obtain data on reported events of “myocarditis” and “pericarditis” amongst immunocompromised individuals following COVID-19 mRNA vaccination (COVID-19 Vaccines Pfizer/BioNTech [Comirnaty] and Moderna [Spikevax]). Searches of the VAERS and EudraVigilance databases covered the period from the date of vaccine launch until 30 November 2021.

Meanwhile, systematic searches were carried out in the United Kingdom’s Yellow Card scheme’s database of spontaneous reports to attain reports of “myocarditis” and “pericarditis” following mRNA vaccination for immunocompromised individuals. These searches were carried out by (Medicines and Healthcare Regulatory Agency (MHRA) staff and supplied to us for analysis. The datalock point was 01 December 2021. Counts of overall reports of myocarditis and pericarditis were obtained within a personal communication from the MHRA. Further details regarding the overall population fatalities were obtained from the ‘Coronavirus vaccine - weekly summary of Yellow Card reporting’ (12). The datalock point for this data was 08 December 2021. Immunocompromised individuals were defined as patients who were: transplant recipients, HIV/AIDs patients, or cancer patients using chemotherapies. Counts of myocarditis and pericarditis reports were tabulated for each immunocompromised subgroup.

### Full details of these searches are provided in the Supplementary file

The level of detail within reports differs between spontaneous reporting databases depending on data collection forms used. In the VAERS database, it is possible to search for comorbidities, while in EudraVigilance it is necessary to use concomitant immunosuppressive treatments as a surrogate for immunosuppression. Therefore, in the VAERS database we searched for reports of myocarditis or pericarditis following at least one dose of COVID-19 mRNA vaccine (COVID-19 Vaccine Pfizer/BioNTech [Comirnaty] or COVID-19 Moderna [Spikevax]), where a history of transplantation, was reported. To align the data analysis both databases were searched for medications which could be used as a surrogate marker for transplantation, HIV/AIDS, or cancer; these medications included anti-rejection treatments, steroids, HIV/AIDS treatments, and anti-cancer therapeutics approved in the US and EU (13-15). A full list of these medications is provided in the Supplementary file. Methotrexate was included as a medication proxy for auto-immune conditions and cancer, however due to its wide use in autoimmune disease methotrexate was treated as a separate category. Age group and sex were obtained from each report and presented in a flowchart. Where reported, time-to-onset and vaccine dose were extracted and tabulated. Seriousness was obtained for each case, as defined in ICH Topic E2A: Clinical Safety Data Management, namely myocarditis and pericarditis events which were fatal, caused or prolonged hospitalisation (including surgery, where specified), or were life-threatening (16). Two researchers independently searched the spontaneous reporting databases and extracted data.

It is not possible to estimate incidence using spontaneously reported data due to inherent limitations of spontaneous reporting systems (underreporting of events and the lack of a denominator in the database). Instead, proportional reporting rates (PRR) were calculated using the method described by Evans et al. (17). Unlike incidence rates which evaluates the number of events relative to the total population exposed, PRR uses the total ADRs reported following exposure to the vaccine in each subgroup as the denominator. A ratio of reports of myocarditis and pericarditis from immunocompromised subgroups is then compared with the total myocarditis and pericarditis reports from all vaccinees following any dose of COVID-19 mRNA vaccination. PRR was calculated for the VAERS population only, due to limitations of the search functions available for the EudraVigilance dataset.

### Patient and Public Involvement

Patients and the public were not consulted during this study due to the data source used. No identifiable information is included within the outputs of spontaneous reporting systems.

## Results

### European Union/European Economic Area

There were 50 individual cases of myocarditis or pericarditis submitted to EudraVigilance up to 30 November 2021 with EU/EEA origin, amongst patients with immunosuppression as suggested by concomitant medications. Myocarditis was detailed in 19 (38.0%) of the 50 total reports, while pericarditis was reported for 29 (58.0%) cases. There were two reports (4.0%) where both myocarditis and pericarditis were described. Most reports were from patients in the 18-64 years age group (59.5% of 50 reports); 92.0% of reports concerned adults aged 18 years or older (Figure 1). Twenty-nine of the 50 reports involved a male patient (58.0%; Figure 1). In total, 82 medications of interest were reported within the 50cases submitted to EudraVigilance up to 30 November 2021 (Table 1). Of all concomitant medications investigated, cancer treatments were most commonly reported (n=50 of 82 medications, 61.0%; Table 1). Ciclosporin was least frequently reported (n=1, 1.2%; Table 1).

**Table 1:** Concomitant medications within spontaneous reports of myocarditis and pericarditis submitted to EudraVigilance for the European Union and European Economic Area only, following COVID-19 mRNA vaccines. Reports of myocarditis or pericarditis were searched in the EudraVigilance database following COVID-19 mRNA vaccines, either COVID-19 Vaccine Moderna (Spikevax) or Pfizer/BioNTech (Comirnaty). Medications were used as a proxy for immunocompromised subgroups: transplant recipients (tacrolimus, mycophenolate mofetil, methotrexate, Prednisolone, ciclosporin and steroids [including prednisolone]), HIV infection, and patients undergoing cancer treatment.

|  | **Tacrolimus** | **Mycophenolate mofetil** | **Methotrexate** | **Ciclosporin** | **Prednisolone** | **HIV treatments** | **Cancer treatments\*\*** | **Total\*** |
| --- | --- | --- | --- | --- | --- | --- | --- | --- |
| **Comirnaty** |  |  |  |  |  |  |  |  |
| Myocarditis | 1 | 2 | 5 | 0 | 6 | 1 | 14 | **29** |
| Pericarditis | 0 | 2 | 7 | 1 | 8 | 1 | 25 | **44** |
| Myopericarditis | 0 | 0 | 0 | 0 | 0 | 0 | 2 | **2** |
| **Spikevax** |  |  |  |  |  |  |  |  |
| Myocarditis | 1 | 2 | 1 | 0 | 0 | 0 | 0 | **4** |
| Pericarditis | 0 | 0 | 1 | 0 | 1 | 0 | 1 | **3** |
| Myopericarditis | 0 | 0 | 0 | 0 | 0 | 0 | 0 | **0** |
| **Total\*** | **2** | **6** | **14** | **1** | **15** | **2** | **42** | **82** |
| *Reports may contain more than one concomitant medication, and/or more than one event |  |  |  |  |  |  |  |  |
| **Hydrocortisone excluded as no indications given within reports and the drug is commonly used to treat other conditions |  |  |  |  |  |  |  |  |

**Figure 1:**
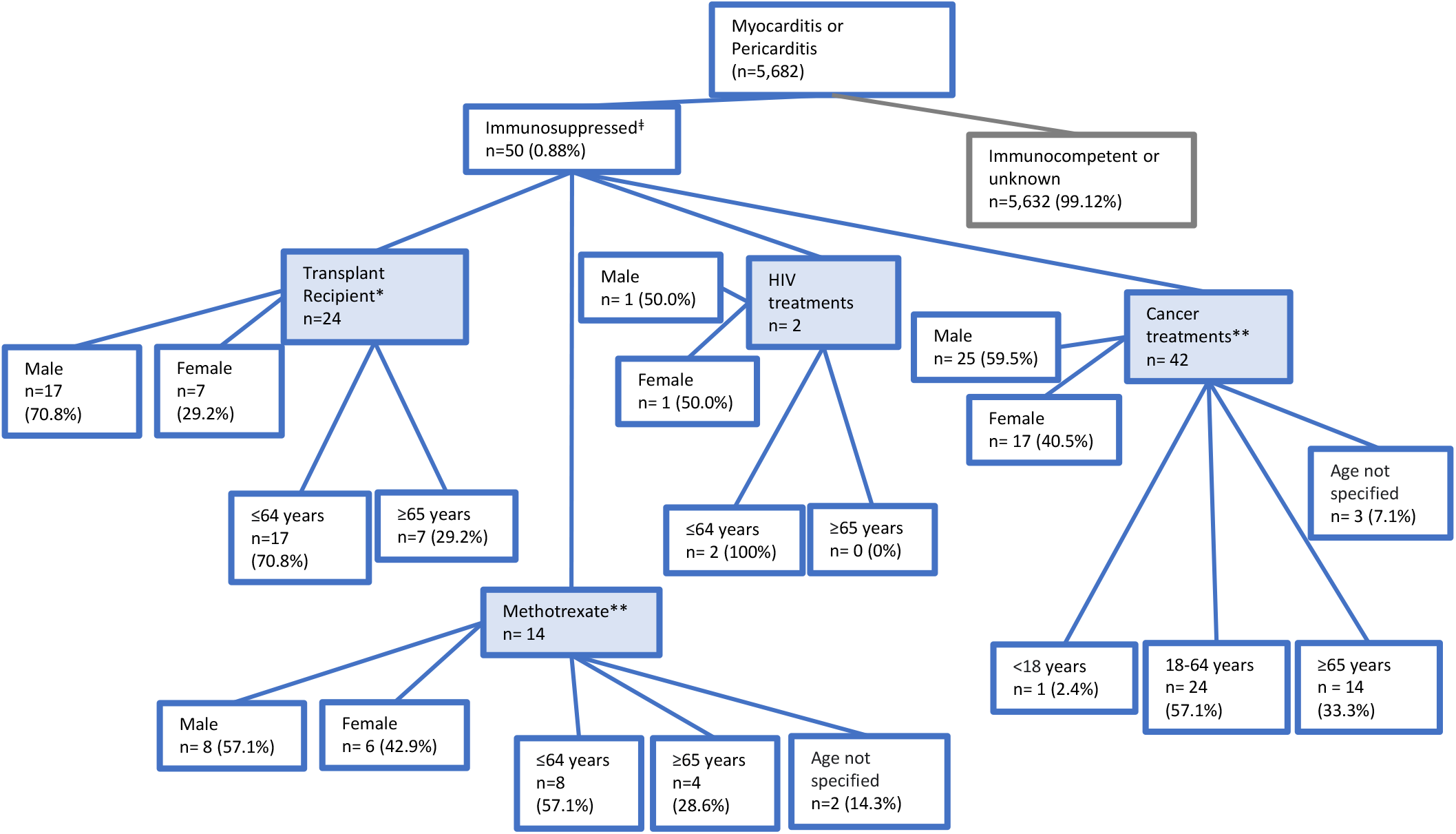
Tree diagram detailing the characteristics of vaccinees submitting spontaneous reports of myocarditis and pericarditis to EudraVigilance for EU and EEA only, following COVID-19 mRNA vaccines in immunocompromised individuals. Reports of myocarditis or pericarditis were searched in the EudraVigilance database following COVID-19 mRNA vaccines, either Moderna (Spikevax) or Pfizer/BioNTech (Comirnaty) (n=50 reports). The resultant reports were further assessed for individuals that were likely to be immunosuppressed based on the following search terms (transplant medications*, HIV/AIDS treatments, EU-approved cancer therapies). Concomitant medications were used as a proxy for disease status. ^≠^Counts are not mutually exclusive; one report may contain multiple concomitant medications from different categories. *Where tacrolimus, mycophenolate mofetil, ciclosporin, or prednisolone were reported as concomitant medications. **Methotrexate was included as a potential transplant medication, however due to its wide use in autoimmune disease this was listed independently of the treatments used as a proxy for transplantation.

Of the 50 reports from immunocompromised individuals, 38 (76.0%) met the criteria for a ‘serious’ event (defined as: event caused or prolonged hospitalisation, was life threatening, or had a fatal outcome; Table 2). None of the myocarditis or pericarditis events reported in immunocompromised subgroups of interest up to 30 November 2021 had a fatal outcome. In contrast, 75.78% of myocarditis or pericarditis events reported to EudraVigilance in the population overall met the criteria for a serious case (n=4309 of 5681 cases reported following a COVID-19 mRNA vaccine, overall). Of these 5681 cases, 156 (2.7%) reported a fatal outcome.

**Table 2:** Breakdown of serious myocarditis and pericarditis events reported via EudraVigilance and VAERS. Reports of myocarditis, pericarditis or myopericarditis were searched in the VAERS database following COVID-19 mRNA vaccines, either Moderna or Pfizer/BioNTech. The resultant reports were further assessed for individuals that were likely to be immunocompromised based on the following search terms (transplant, medicinal immunosuppressant drugs; tacrolimus, mycophenolate mofetil, methotrexate, Prednisolone, ciclosporin and Steroids, HIV or approved cancer therapeutics). Serious events were determined as required hospitalisation, surgery or fatality as detailed in the comments box. Values indicate the events recorded and the percentage of the total events in each of the four characteristic categories with a 95% confidence interval.

| Immunocompromised<br>Characteristic | Serious Events<br>EudraVigilance* |  |  | Serious events<br>VAERS |  |  | Total |  |  |
| --- | --- | --- | --- | --- | --- | --- | --- | --- | --- |
|  | n | %<sup>‡</sup> (95% CI) | Total<br>reported<br>in<br>subgroup | n | %<sup>‡</sup> (95% CI) | Total<br>reported<br>in<br>subgroup | n | %<sup>‡</sup> (95% CI) | Total<br>reported<br>in<br>subgroup |
| Transplant | 22 | 91.7 (73.0-99.0) | **24** | 2 | 20.0 (2.5-55.6) | **10** | 24 | 70.5 (52.5-84.9) | **34** |
| Methotrexate-treated patients | 11 | 78.6 (49.2-95.3) | **14** | 5 | 55.6 (21.2-86.3) | **9** | 16 | 69.6 (47.1-86.8) | **23** |
| HIV | 2 | 100 | **2** | 3 | 42.9 (9.9-81.6) | **7** | 5 | 55.5 (21.2-86.3) | **9** |
| Cancer Therapeutics | 32 | 65.3 (50.4-78.3) | **49** | 13 | 41.9 (24.5-60.9) | **31** | 45 | 56.3 (44.7-67.3) | **80** |
| CI = Confidence Interval<br>*Counts not mutually exclusive, multiple medications could be reported in the same case<br>‡Percentage of total reports within the EudraVigilance or VAERS database for that subgroup<br>NB: Serious event defined as: event caused or prolonged hospitalisation, required surgery, was life threatening, or was fatal |  |  |  |  |  |  |  |  |  |

### United States

Meanwhile in the VAERS spontaneous reporting system, there was a total of 3,062 reports of myocarditis or pericarditis following mRNA vaccination (Pfizer or Moderna), of which 1.83% (n=57; Figure 2) were from immunocompromised individuals. These spontaneous reports of myocarditis or pericarditis in immunocompromised individuals differed in their gender and age distribution compared to the whole population with myocarditis or pericarditis (18). The whole population had a bias of younger males experiencing myocarditis or pericarditis following COVID-19 mRNA vaccinations (18), whereas from the immunocompromised population 52.6% of these events occurred in males and 50.9% were under 60 years of age. Thus, this could indicate a broadening in those susceptible to myocarditis or pericarditis in immunocompromised individuals. Splitting the analysis of immunocompromised individuals into transplant recipients, medically induced immunosuppression using methotrexate, HIV, and cancer is indicated in Figure 2. Within these four subsets we investigated whether there were patterns in time-to-onset or vaccination dose for reported myocarditis or pericarditis events (Table 3). Predominately events were reported after the second mRNA vaccination (mean 51.8%; range 33.3 – 61.3% within immunocompromised subgroups), while 8.9% of events (range 0 – 16.1% within immunocompromised subgroups) were reported following the third vaccination dose (Table 3). Approximately 70% of reported events occurred within 14 days of vaccination (n=39; range 44.4 – 100% within immunocompromised subgroups; Table 3). Therefore, this is suggestive of a potential causal effect due to the rapid onset of symptoms following exposure to a COVID-19 mRNA vaccine. The seriousness of each event, (resulting in hospitalisation, surgery, or fatality) was assessed and recorded in Table 2, with 41.1% (n=23) of the 56 reports of myocarditis or pericarditis from immunocompromised subgroups meeting the criteria for ‘serious.’ Surgery was required for three individuals, and one fatality was recorded. The fatal event occurred in a female undergoing cancer therapy, eight days after receiving the first dose of COVID-19 Vaccine Moderna. Cause of death was not specified within the report, however an autopsy demonstrated systemic inflammatory reaction, which affected primarily the small blood vessels of the brain with leptomeninges, heart, lungs, and liver also affected. Generalised systemic tissue inflammation caused diffuse intravascular inflammatory microthrombi and haemorrhagic myocarditis. In comparison, for all myocarditis and pericarditis reports 63.4% were classified as serious, this includes immunocompromised as well as immune-competent individuals.

**Table 3:** Spontaneous reports of myocarditis and pericarditis on the VAERS database following COVID-19 mRNA vaccines in immunocompromised individuals subdivided into time to onset and vaccine dosage. Reports of myocarditis, pericarditis or myopericarditis were searched in the VAERS database following COVID-19 mRNA vaccines, either Moderna or Pfizer/BioNTech. The resultant reports were further assessed for individuals that were likely to be immunocompromised based on the following search terms (transplant, immunosuppressant drugs; tacrolimus, mycophenolate mofetil, methotrexate, Prednisolone, ciclosporin and Steroids, HIV or approved cancer therapeutics: https://www.cancer.gov/about-cancer/treatment/drugs). Values indicate the events recorded and as a percentage of the total in each of the four characteristic categories with 95% confidence intervals.

| Reports to VAERS |  |  |  |  |  |  |  |  |  |  |  |  |  |  |  |
| --- | --- | --- | --- | --- | --- | --- | --- | --- | --- | --- | --- | --- | --- | --- | --- |
| Immuno-compromised characteristic | Outcome |  |  | Vaccine dose |  |  |  |  |  |  |  | Time to Onset of Symptoms |  |  |  |
|  |  | n | % (95 % CI) | Dose 1 |  | Dose 2 |  | Dose 3 |  | Unknown dose |  | <14 days |  | >14 days |  |
|  |  |  |  | n | % (95% CI) | n | % (95% CI) | n | % (95% CI) | n | % (95% CI) | n | % (95% CI) | n | % (95% CI) |
| Transplant | Myocarditis | 1 | 11.1 (0.3-48.2 ) | 0 | 0.00 | 2 | 22.2 (2.8-60.0) | 0 | 0.0 | 0 | 0.0 | 0 | 0.0 | 2 | 22.2 (2.8-60.0) |
|  | Pericarditis | 5 | 55.6 (21.2-86.3 ) | 1 | 11.1 (0.3-48.2) | 2 | 22.2 (2.8-60.0) | 0 | 0.0 | 3 | 33.3 | 4 | 44.4 (13.7-78.8) | 2 | 22.2 (2.8-60.0) |
|  | Myopericar ditis | 3 | 33.3 (7.5-70.1 ) | 1 | 11.1 (0.3-48.2) | 1 | 11.1 (0.3-48.2) | 0 | 0.0 | 0 | 0.0 | 1 | 11.1 (0.3-48.2) | 1 | 11.1 (0.3-48.2) |
|  | Total | 9 | 100.0 | 3 | 33.3 (7.5-70.7) | 3 | 33.3 (7.5-70.7) | 0 | 0.0 | 3 | 33.3 (7.5-70.7) | 4 | 44.4 (13.7-78.8) | 5 | 55.6 (21.2-86.3) |
| Methotrexate | Myocarditis | 0 | 0.0 | 0 | 0.0 | 0 | 0.0 | 0 | 0.0 | 0 | 0.0 | 0 | 0.0 | 0 | 0.0 |
|  | Pericarditis | 8 | 88.9 (51.8-99.7) | 4 | 44.4 (13.7-78.8) | 4 | 44.4 (13.7-78.8) | 0 | 0.0 | 0 | 0.0 | 6 | 66.7 (29.9-92.5) | 1 | 11.1 (0.3-48.2) |
|  | Myopericarditis | 1 | 11.1 (0.3-48.2) | 0 | 0.0 | 0 | 0.0 | 0 | 0.0 | 1 | 11.1 (0.3-48.2) | 1 | 11.1 (0.3-48.2) | 0 | 0.0 |
|  | Total | 9 | 100.0 | 4 | 44.4 (13.7-78.8) | 4 | 44.4 (13.7-78.8) | 0 | 0.0 | 1 | 11.1 (0.3-48.2) | 7 | 77.8 (40.0-97.2) | 1 | 11.1 (0.3-48.2) |
| HIV | Myocarditis | 2 | 28.6 (3.7-71.0) | 1 | 14.3 (0.4-57.9) | 1 | 14.3 (0.4-57.9) | 0 | 0.0 | 0 | 0.0 | 2 | 28.6 | 0 | 0.0 |
|  | Pericarditis | 4 | 57.1 (18.4-90.1) | 3 | 42.9 (9.9-81.6) | 1 | 14.3 (0.4-57.9) | 0 | 0.0 | 0 | 0.0 | 4 | 57.1 (18.4-90.1) | 0 | 0.0 |
|  | Myopericarditis | 1 | 14.3 (0.4-57.9) | 0 | 0.0 | 1 | 14.3 (0.4-57.9) | 0 | 0.0 | 0 | 0.0 | 1 | 14.3 (0.4-57.9) | 0 | 0.0 |
|  | Total | 7 | 100.0 | 4 | 57.1 (18.4-90.1) | 3 | 42.9 (9.9-81.6) | 0 | 0.0 | 0 | 0.0 | 7 | 100.0 | 0 | 0.0 |
| **Cancer Therapeutics** | Myocarditis | 10 | 32.3<br>(16.7-51.4) | 2 | 6.5 (0.8-21.4) | 6 | 19.4<br>(7.5-37.5) | 1 | 3.2 (0.1-16.7) | 1 | 3.2 (0.1-16.7) | 8 | 25.8 (11.9-44.6) | 2 | 6.5 (0.8-21.4) |
|  | Pericarditis | 17 | 54.8<br>(36.0-72.7) | 4*<br>* | 12.9 (3.6-29.8) | 11 | 35.5<br>(19.2-54.6) | 2 | 6.5 (0.8-21.4) | 0 | 0.0 | 10*<br>* | 32.3 (16.7-51.4) | 7 | 22.6 (9.6-41.1) |
|  | Myopericarditis | 4 | 12.9<br>(3.6-29.8) | 0 | 0.0 | 2 | 6.5<br>(0.8-21.4) | 2 | 6.5 (0.8-21.4) | 0 | 0.0 | 3 | 9.7 (2.0-25.8) | 0 | 0.0 |
|  | Total | 31 | 100.0 | 6 | 19.4 (7.5-37.5) | 19 | 61.3<br>(42.2-78.2) | 5 | 16.1 (5.5-33.7) | 1 | 3.2 (0.1-16.7) | 21 | 67.7 (48.6-83.3) | 9 | 29.0 (14.2-48.0) |
| **All** | Total | 56 | 100.0 | 17 | 28.1 (18.8-44.1) | 31 | 55.4<br>(41.5-68.7) | 5 | 8.9 (3.0-19.6) | 5 | 8.9 (3.0-19.6) | 39 | 69.6 (55.9-81.2) | 15 | 26.8 (15.8-40.3) |
| * n=1, unspecified time to onset<br>**n=1, fatality<br>CI=Confidence Interval |  |  |  |  |  |  |  |  |  |  |  |  |  |  |  |

**Figure 2:**
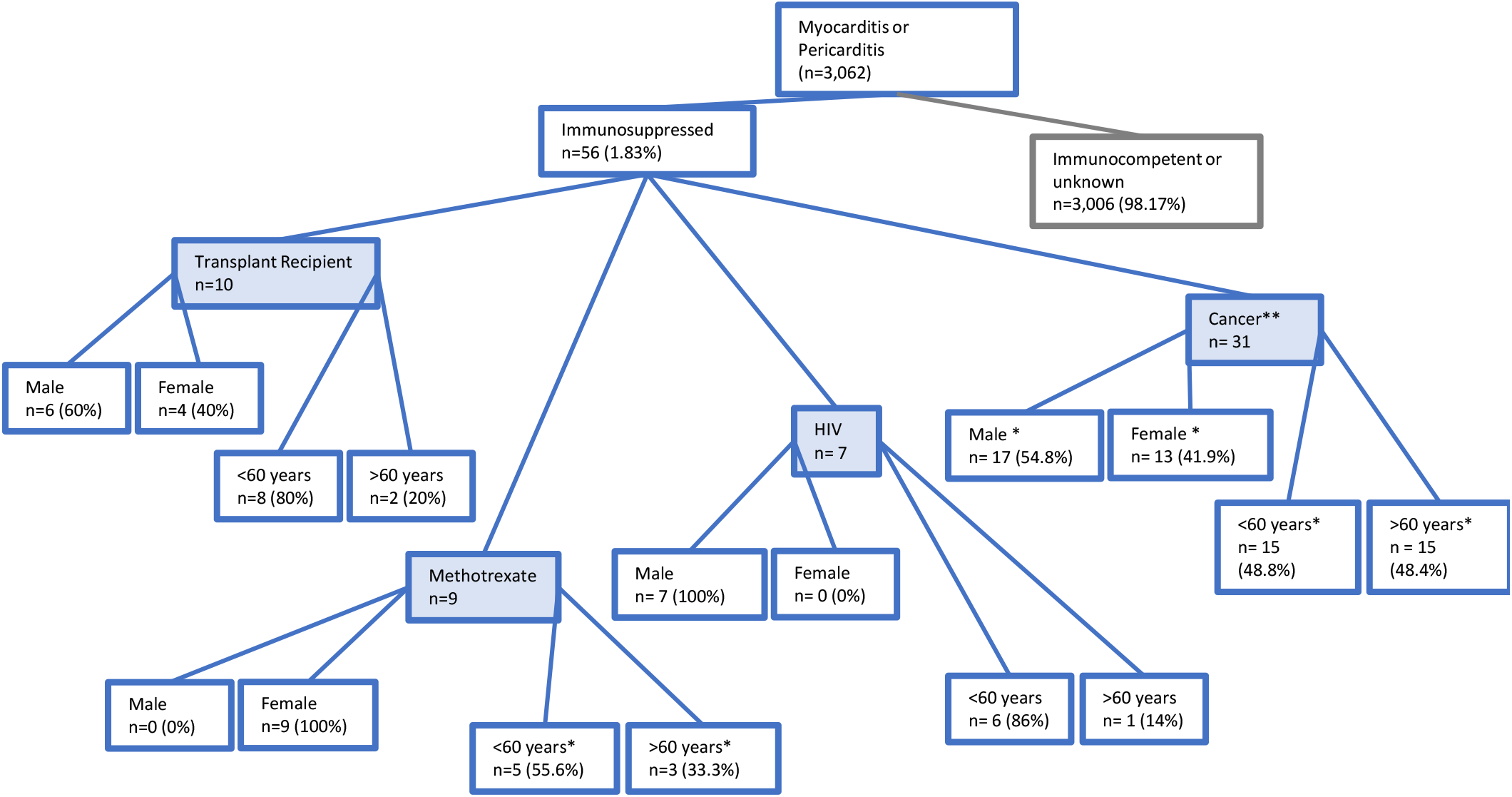
Tree diagram detailing the characteristics of spontaneous reports of myocarditis and pericarditis on the VAERS database following COVID-19 mRNA vaccines in immunocompromised individuals. Reports of myocarditis, pericarditis or myopericarditis were searched in the VAERS database following COVID-19 mRNA vaccines, either Moderna or Pfizer BioNTech. The resultant reports were further assessed for individuals that were likely to be immunocompromised based on the following search terms (transplant, tacrolimus, mycophenolate mofetil, methotrexate, prednisolone, ciclosporin). Transplant recipients were assessed separately to immunocompromised individuals that received medication, independent of disease setting. * unspecified category, ** cancer defined by use of cancer approved medicines

To determine whether these events were reported at higher levels in immunocompromised compared to immune-competent (or unspecified) individuals a PRR was calculated. This was only possible in the VAERS dataset, due to data limitations in the EudraVigilance and Yellow Card datasets. In the VAERS population, the reporting rate of myocarditis and pericarditis was slightly higher for immunocompromised patients (transplant recipients, HIV/AIDS patients, and cancer patients) compared with immune competent individuals (PRR=1.36 [95% CI: 0.89-1.82]). However, no statistical differences were observed. It should be noted that PRR are not the same as incidence rates. Data on duration of symptoms was not available for either database examined.

### United Kingdom

In total, 1009 reports of myocarditis and pericarditis following either COVID-19 mRNA vaccination had been submitted to the Yellow Card scheme up to 01 December 2021 (12). As of 08 December 2021, there had been three reported to have a fatal outcome; all three fatal events were in people who had received Pfizer/BioNTech (12). The reporting rate for Pfizer/BioNTech was 11 cases of myocarditis reported per million, and eight cases of pericarditis reported per million first or second vaccine doses. For Moderna, there were 39 reports of myocarditis doses and 22 reports of pericarditis per million first and second doses of the vaccine administered (12). Reporting rates were highest for the 19-29 years age group for both COVID-19 mRNA vaccines, with a trend for decreased reporting in older age groups in the population overall (12).

Amongst immunocompromised individuals overall, there were 72 cases of myocarditis, pericarditis, or myopericarditis reported to the Yellow Card scheme up to 01 December 2021 (Figure 3). Therefore, of all myocarditis or pericarditis reports recorded in the UK, 7.1% were from immunocompromised individuals. The majority of myocarditis and pericarditis events were from people aged <65 years, similar to the overall population who had reported events in the UK, whereas the male dominance of events seen in the whole population (74.9% of myocarditis and pericarditis were from males) (18) was not seen in the immunocompromised, here 51.4% of reports were from females (n=37; Figure 3). Further classification of these events reported in the UK are detailed in Table 4. One event was excluded from the analysis as it was not classified as myocarditis or pericarditis, rather it described general cardiac issues following COVID-19 mRNA vaccination.

**Table 4:** Spontaneous reports of myocarditis and pericarditis submitted to the Yellow Card Scheme in the UK following COVID-19 mRNA vaccines in immunocompromised individuals. Reports of myocarditis or pericarditis were searched in the Yellow Card scheme following COVID-19 mRNA vaccines, either COVID-19 Vaccine Moderna (Spikevax) or Pfizer/BioNTech (Comirnaty). Events were classified by vaccination dose and time to onset of symptoms. *n=4 missing information on time-to onset, **n=1 excluded due to not classification by myocarditis, pericarditis or myopericarditis.

| Reports to Yellow Card (UK) |  |  |  |  |  |  |  |  |  |  |  |  |  |  |
| --- | --- | --- | --- | --- | --- | --- | --- | --- | --- | --- | --- | --- | --- | --- |
| Outcome |  |  | Vaccine Dose |  |  |  |  |  |  |  | Time to Onset of Symptoms |  |  |  |
|  |  |  | Dose 1 |  | Dose 2 |  | Dose 3 |  | Unknown |  | <14 days |  | >14 days |  |
|  | n | % (95% CI) | n | % (95% CI) | n | % (95% CI) | n | % (95% CI) | n | % (95% CI) | n | % (95% CI) | n | % (95% CI) |
| **Myocarditis** | 30 | 42.3 (30.6-54.6) | 13 | 18.3 (10.1-29.3) | 11 | 15.5 (8.0-26.0) | 5 | 7.0 (2.3-15.7) | 1 | 1.4 (0.0-7.6) | 19 | 26.8 (16.9-38.6) | 8 | 11.3 (5.0-21.0) |
| **Pericarditis** | 38 | 53.5 (41.3-65.5) | 9 | 12.7 (6.0-22.7) | 14 | 19.7 (11.2-30.9) | 12 | 16.9 (9.0-27.7) | 3 | 4.2 (0.9-11.9) | 24 | 33.8 (23.0-46.0) | 13 | 18.3 (10.1-29.3) |
| **Myopericarditis** | 3 | 4.2 (0.9-11.9) | 0 | 0 | 3 | 4.2 (0.9-11.9) | 0 | 0 | 0 | 0 | 2 | 2.8 (0.3-9.8) | 1 | 1.4 (0.0-7.6) |
| **Total** | 71** | 100 | 22 | 31.0 (20.5-43.1) | 28 | 39.4 (28.0-51.7) | 17 | 23.9 (14.6-35.5) | 4 | 5.6 (1.6-13.8) | 45 | 63.4 (51.1-74.5) | 22 | 31.0 (20.5-43.1) |
| CI=Confidence Interval |  |  |  |  |  |  |  |  |  |  |  |  |  |  |

**Figure 3:**
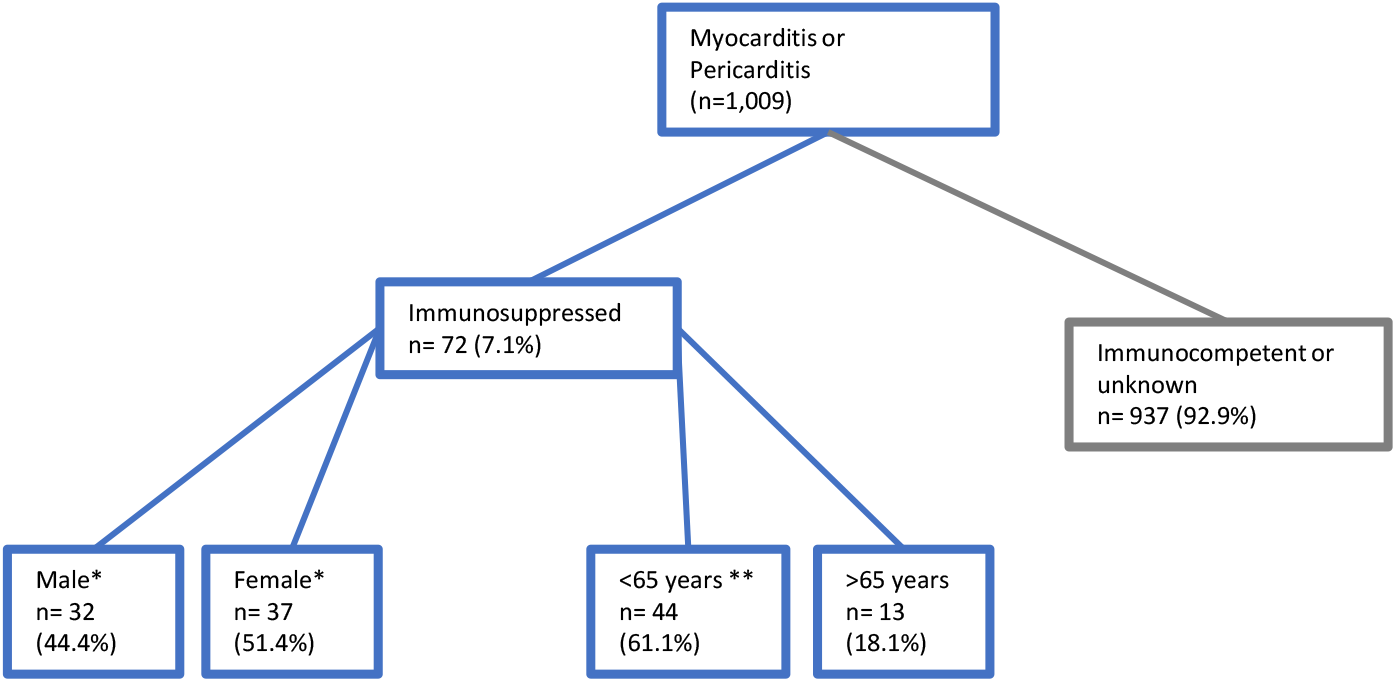
Tree diagram detailing the characteristics of spontaneous reports of myocarditis and pericarditis on the Yellow Card database following COVID-19 mRNA vaccines in immunocompromised individuals (Yellow Card, UK) Reports of myocarditis, pericarditis or myopericarditis from immunocompromised individuals were searched in the Yellow Card scheme following COVID-19 mRNA vaccines, either Moderna or Pfizer BioNTech. Data provided by MRHA, datalock 1^st^ December 2021. * n=3 unspecified, ** n=2 under 18 years. For age category n=5 unknown.

### Myocarditis and pericarditis may occur more often in cancer patients following Pfizer mRNA vaccination

Next, we determined whether myocarditis and pericarditis in the immunocompromised subgroups, categorised as: transplant recipients, methotrexate-treated patients, HIV patients or individuals on approved cancer therapeutics (please see supplementary file for full medication lists) differed with the vaccines manufacturer used. This analysis was undertaken using the EudraVigilance and VAERS datasets only, due to how the information was reported from these systems allowed for stratification in this manner. Of the reports in these two datasets Pfizer/BioNTech had a higher number of reports, although details on total vaccines, and from which manufacturer, given in these discrete immunocompromised populations is unknown, which may have impacted the higher number of reports following Pfizer/BioNTech vaccine administration compared to Moderna. Vaccine roll out, vaccine interval period and demographics of recipients of each vaccine in each region were unknown and may have differed, which may have led to higher reports following Pfizer/BioNTech administration compared to Moderna. This is inconsistent with the overall reporting population where higher reporting rates from Moderna were seen, potentially due to higher amounts of mRNA contained with Moderna vaccines compared to Pfizer/BioNTech (100 µg vs 30 µg, respectively) (19, 20). To determine whether the number of reports from each immunocompromised subgroup differed following each vaccine type, the percentage of reports was calculated using the total number of myocarditis and pericarditis reports from each vaccine type as the denominator. Following Pfizer/BioNTech vaccine administration 57.8% of reports of myocarditis and pericarditis were received from individuals on cancer therapeutics, whereas a lower percentage of reports was received following Moderna vaccination (Table 5). This data is suggestive that potentially different immunocompromised conditions may require careful consideration as to the type of mRNA COVID-19 vaccines given.

**Table 5:** Myocarditis and pericarditis adverse events reported following Moderna or Pfizer/BioNTech mRNA vaccination stratified by immunocompromised sub-groups. Reports of myocarditis, pericarditis or myopericarditis were searched in the EudraVigilance and VAERS database following COVID-19 mRNA vaccines, either Moderna or Pfizer/BioNTech. The resultant reports were further assessed for individuals that were likely to be immunocompromised based on the following search terms (transplant, immunosuppressant drugs; tacrolimus, mycophenolate mofetil, methotrexate, Prednisolone, ciclosporin and Steroids, HIV or approved cancer therapeutics: https://www.cancer.gov/about-cancer/treatment/drugs). Values indicate the events reported from each vaccine manufacture within each immunocompromised category. Percentage of the total reports for each manufacture are calculated for each of the characteristic categories

| Immunocompromised<br>Characteristic | EudraVigilance |  | VAERS |  | Total |  |  |  |
| --- | --- | --- | --- | --- | --- | --- | --- | --- |
|  | Pfizer/BioNTech | Moderna | Pfizer/BioNTech | Moderna | Pfizer/BioNTech |  | Moderna |  |
|  |  |  |  |  | n | % (95% CI) | n | % (95% CI) |
| **Transplant** | 20 | 4 | 7 | 2 | 27 | 24.77 (17.00-33.96) | 6 | 20.69 (7.99-39.72) |
| **Methotrexate-treated patients** | 12 | 2 | 1 | 8 | 13 | 11.93 (6.51-19.53) | 10 | 34.48 (17.94-54.33) |
| **HIV** | 2 | 0 | 4 | 3 | 6 | 5.5 (2.05-11.60) | 3 | 10.34 (2.19-27.35) |
| **Cancer Therapeutics** | 41 | 1 | 22 | 9 | 63 | 57.8 (47.96-67.20) | 10 | 34.48 (17.94-54.33) |
| **Total** | 75 | 7 | 34 | 22 | 109 | 100 | 29 | 100 |
| CI=Confidence Interval |  |  |  |  |  |  |  |  |

### Myocarditis and pericarditis events are not restricted to mRNA based COVID-19 vaccines

Vaccine-associated myocarditis and pericarditis following COVID-19 vaccination is not only restricted to the use of mRNA-based vaccinations. Reports of myocarditis and pericarditis have been reported following adenovirus vector COVID-19 vaccines, suggesting it may not be the mRNA component of vaccines which trigger this response but the immune response or elements of SARS-CoV-2 itself. In the UK, 385 events of myocarditis and pericarditis had been reported via the Yellow Card scheme following administration of the AstraZeneca adenovirus vector COVID-19 vaccine, up to 15 December 2021 (12), which includes immunocompromised as well as immunocompetent populations. Meanwhile a total of 348 events have been reported to EudraVigilance following vaccination with AstraZeneca or Janssen COVID-19 vaccines (Table 6) and 141 events following Janssen vaccination have been reported to VAERS (Table 6). We were able to stratify the events reported to EudraVigilance and VAERS into those reported by immunocompromised individuals and immunocompetent and calculated the percentage of total events from each dataset that were associated with these two population groups (Table 6). The number of myocarditis and pericarditis events is much lower following viral vector vaccines, although direct comparison is not advisable due to differing total number of vaccinees receiving each type of vaccine. Percentages of reports from immunocompromised individuals are comparable for mRNA-based vaccines and adenovirus vector vaccines, with 95% confidence intervals which overlap (Table 6).

**Table 6:** Comparison of the number of myocarditis and pericarditis adverse events following the different COVID-19 vaccination types. Reports of myocarditis, pericarditis or myopericarditis were searched in the EudraVigilance and VAERS database following COVID-19 mRNA vaccines, either Moderna or Pfizer/BioNTech, and adenovirus vector vaccines (AstraZeneca or Janssen). Comparison was made between the immunocompromised group identified in our analysis and the overall reporting population to the EudraVigilance and VAERS databases for these vaccines.

|  |  | mRNA vaccines |  | Adenovirus vector vaccines |  |
| --- | --- | --- | --- | --- | --- |
|  |  | Events (n) | % (95% CI)* | Events (n) | % (95% CI)* |
| **EudraVigilance** | Immunocompromised | 50 | 0.9 (0.7-1.2) | 5 | 1.43 (0.47-3.32) |
|  | Immunocompetent or unknown | 5632 | 99.1 (98.8-99.3) | 343 | 98.56 (96.68-99.53) |
|  | **Total** | **5682** | 100.0 | **348** | 100.0 |
| **VAERS** | Immunocompromised | 57 | 1.86 (1.41-2.40) | 3 | 2.13 (0.44-6.09) |
|  | Immunocompetent or unknown | 3006 | 98.14 (97.60-98.59) | 138 | 97.87 (93.91-99.56) |
|  | **Total** | **3063** | 100.0 | **141** | 100.0 |
| *Percentage of total |  |  |  |  |  |

### Mechanisms of myocarditis and pericarditis following mRNA COVID-19 vaccination

The mechanisms for myocarditis or pericarditis following COVID-19 mRNA vaccination are not yet fully understood. It has not yet been determined whether it is the mRNA element itself or the ‘self’ generated COVID-19 spike protein from the mRNA message within the vaccines or the resultant immune response, which cardiac tissues are sensitive to.

Following infection or vaccination, the adaptive immune system is activated. Adaptive immunity involves lymphocytes, particularly B-cells and certain T cells, that bind and recognise the foreign antigen. Upon first infection, or vaccination, the pathogen is bound by naïve B cells via the B cell receptor (BCR) or processed by antigen presenting cells (APCs) into smaller fragments, known as antigens. In order to mount an appropriate immune response, B cells will undergo differentiation and maturation within the lymph nodes in the germinal centre reaction (Figure 4). The germinal centre reaction results in the generation of high affinity, high avidity BCRs for the antigen and leads to the production of antibody producing/secreting plasma B cells and memory B cells (Figure 4). Memory B cells will retain the ability to recognise this antigen after the infection has been cleared from the body and is able to rapidly produce plasma B cells and antibodies should the body encounter the pathogen again, rapidly eliminating a second challenge of infection. Simultaneously, T cells will undergo maturation in the thymus in the presence of foreign antigen presented by APCs and through the process of positive and negative selection results in deletion of T cells that bind with high avidity to ‘self’ molecules and promotes expansion of T cells that bind to the foreign antigen. This process of positive and negative selection also gives rise to immune tolerance where there is some cross-over in recognition of self-antigens (21). This immune-tolerance process may give rise to T cells that are reactive to self-antigens, in particular heart auto-reactivity (22), that could lead to myocarditis and pericarditis.

**Figure 4:**
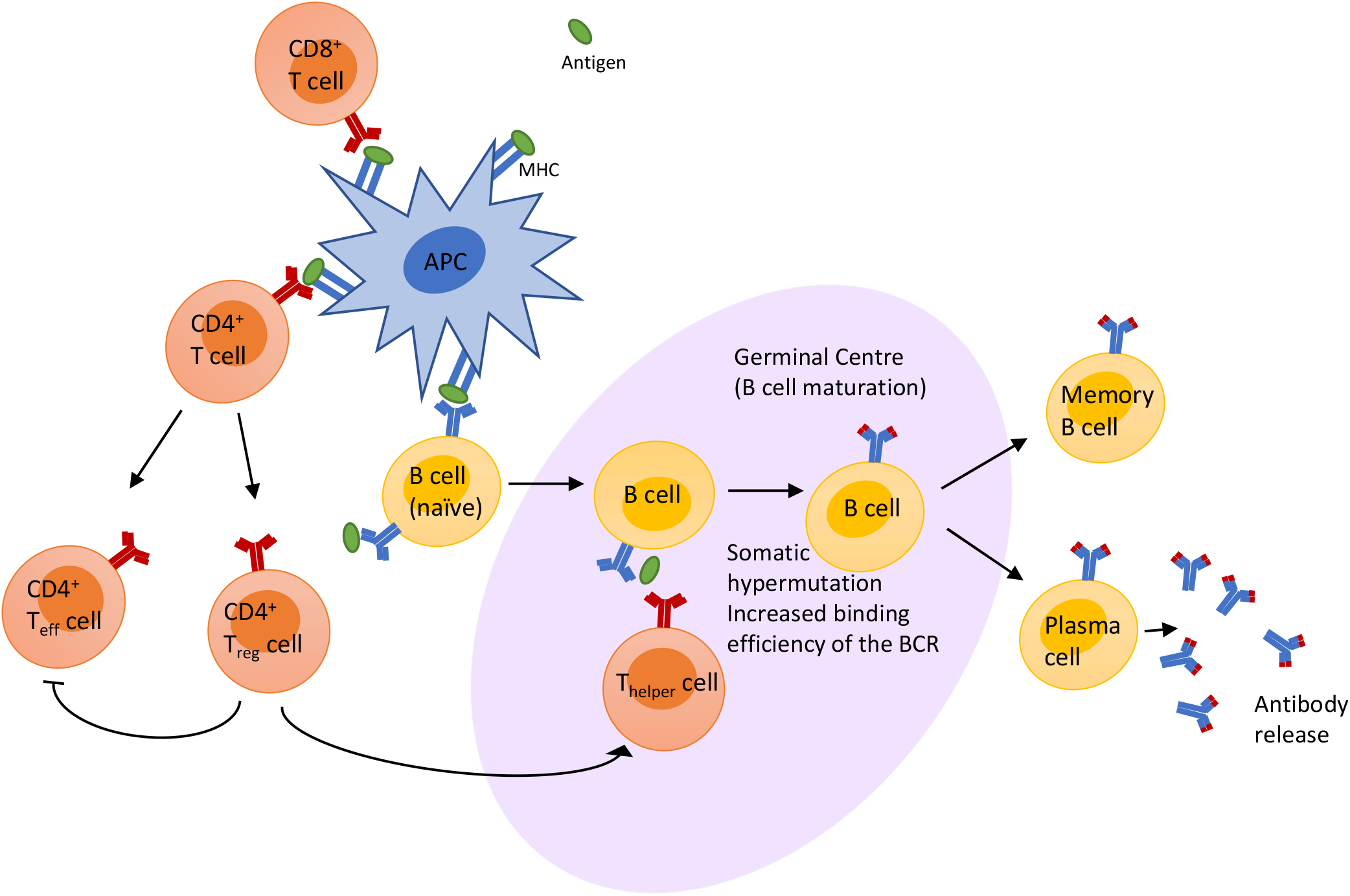
Schematic of the immune response to antigen. Antigen is part of a foreign/non-self pathogen and is presented by an antigen presenting cell (APC) either following viral protein digestion and presentation on the major histocompatibility complex (MHC) or via mRNA translation of the protein within the APC then the protein is presented on the MHC. The non-self protein presented on the MHC is recognised by B and T cells. This triggers a cascade of intracellular signalling responses and leads to the maturation of the B cells, within the germinal centre of a lymph node. The germinal centre reaction involves interactions with B cells, antigen and T cells; leading to somatic hypermutation of the B cell receptor (BCR), generating a BCR that is better adapted to recognise the antigen. The B cell then differentiates into memory B cells and plasma B cells to release the antibodies into the general circulation to clear the body of the infection.

There are multiple hypotheses suggested by Das et al (23) that may explain the mechanism of myocarditis and pericarditis in response to the COVID-19 mRNA vaccines, including;

1. “*mRNA vaccines might generate very high antibody response producing a response similar to multisystem inflammatory syndrome in children (MIS-C) that is associated with COVID infection;*
2. *induction of anti-idiotype cross-reactive antibody-mediated cytokine expression in the myocardium, resulting in aberrant apoptosis and inflammation*,
3. *mRNA vaccines can induce a non-specific innate immune response or a molecular mimicry mechanism may occur, or*
4. *the mRNA itself may be a potent immunogen producing an adjuvant effect*.”

The advantage of mRNA vaccines is their ability to elicit a greater memory response compared to adenovirus vector vaccines (24). The longevity of mRNA vaccines (as evidenced by the persistence of the mRNA and mRNA translation APCs/dendritic cells (DCs) at the injection site and draining lymph nodes) for up to 10 days may enable fine-tuning of the immune responses due to somatic hypermutation, increasing the affinity of the BCR to the immunogen, as well as enabling memory B cell maturation (25) (Figure 4). The ability of mRNA vaccines to produce the immunogen of interest via mRNA translation from within APCs was thought to mimic viral infection, which would elicit a CD8+ cytotoxic T cell response but this is not seen with the COVID-19 mRNA vaccines or in mRNA vaccine trials against rabies virus glycoprotein (RABV-G) or Influenza (H1N1) (25). These mRNA vaccines have demonstrated that, in non-human primates, CD4+ helper T cell responses are elicited (26, 27).

The role of CD4+ T cells in myocarditis has been reviewed by Vdovenko and Eriksson that indicates that CD4+ T cells are the main drivers of heart-specific autoimmunity (28). Therefore, the CD4+ T cell response following mRNA vaccines may trigger myocarditis. CD4+ T cells differentiate into effector or regulatory T cells (T_eff_ or T_reg_, respectively). T_regs_ are vital in immune tolerance to suppress autoimmune T_eff_ cells. Gender differences in circulating T_regs_ has been linked with gender differences in myocarditis (22). Thus the involvement in CD4+ T cells in myocarditis development may indicate that autoimmune responses may occur following mRNA vaccination, via induction of CD4+ T cell expression and may point to the observed differences in myocarditis incidence between males and females following vaccination (18).

Whether the mRNA within the vaccine itself is immunogenic will likely depend on multiple variables within the mRNA sequence. The COVID-19 mRNA vaccines deliver a single stranded mRNA (ssRNA) encoding the spike protein, with modified nucleotides. The modified N^1^-methyl-psuedouridine are known to dampen innate immune responses, while increasing the efficiency of translation *in vivo* (27). As ssRNA is produced by all cells within the body, ssRNA would be recognised as ‘self’ and would not trigger an immune response. Double-stranded RNA (dsRNA) is generally recognised by the cell as viral or is triggered for degradation via the RNA-induced silencing complex (RISC), the cells’ innate ability to modify protein expression by degradation of mRNA (29). Thus the sequence of the ssRNA in the mRNA vaccine will determine the likelihood of complementarity (to itself, other mRNAs or microRNAs) to generate dsRNA which could then trigger immune responses via Toll-like Receptors (TLRs) (30) as well as alter mRNA translation mechanisms (29). Further investigation is needed to determine whether the mRNA sequence is complementary to any ‘self’ mRNA or microRNAs, as this could provide a greater understanding on why certain organs may be preferentially targeted in response to COVID-19 mRNA vaccination. The ability for mRNA vaccines to produce binding and neutralising antibodies in all participants in phase I/II trials (24) demonstrates the ability for these vaccines to activate the immune system similar to viral infection but with high antibody titres (31). Although it should be noted that immunocompromised individuals elicit a reduced antibody response following vaccination (32, 33). Thus, these highly immunogenic mRNA vaccines are activating the B and T lymphocytes, with the potential that heart-specific autoreactive CD4+ T cells may also be activated (or not adequately suppressed correctly via immune-tolerance interactions of T_reg’s_ with T_eff_ cells) potentially leading to reporting of myocarditis and pericarditis (4, 34, 35).

## Discussion and conclusions

Based on the results of spontaneous reporting analyses, there was limited evidence supporting our hypothesis that frequency or seriousness of the myocarditis and pericarditis events reported to spontaneous reporting systems was substantially different for immunocompromised people compared with the reporting population as a whole. In the overall database populations myocarditis and pericarditis seemed to more frequently affect younger males, however the characteristics of those susceptible may have broadened in the immunocompromised population in terms of age and sex; therefore, risk factors for these events are less clear within the immunocompromised population.

Recent reporting of myocarditis and pericarditis in immunocompromised individuals (people on immunosuppressant medications following transplantation, treatment of cancer, or people with HIV infection) raises an important question into how the immune system is responding to mRNA vaccination in these individuals, and whether alternative COVID-19 vaccinations should be considered. Given the current understanding that COVID-19 mRNA vaccines elicit a CD4+ T cell response and the fact that immunocompromised patients have inhibited B and T lymphocyte activity, immune suppression could be a factor which increases the risk of adverse events following COVID-19 mRNA vaccination. In particular, myocarditis and pericarditis following COVID-19 mRNA vaccination may be due to the involvement of CD4+ T cells. Although HIV patients have demonstrated a similar immune response to vaccination compared with healthy adults, other immunocompromised patient subgroups, including those with solid or haematological cancers, solid organ or bone marrow transplantation, and other immunosuppressive conditions, have been shown to generate a lower immune response to COVID-19 vaccination compared with healthy controls (36-39). Therefore, it is possible that immunosuppression due to medication or a medical condition may have an impact on the safety and efficacy of mRNA vaccines against SARS-CoV-2 and other conditions.

Our analysis has not demonstrated a higher frequency of reports of myocarditis and pericarditis amongst the immunocompromised patient subgroups examined (PRR for the VAERS population=1.36 [95% CI: 0.89-1.82]), although the clinical course may be different for these patients compared with immune-competent individuals and requires further monitoring. When the signal first emerged, myocarditis and pericarditis following COVID-19 mRNA vaccination were considered mild conditions of short duration which mostly affected younger males (18, 40-43). Our analysis of reports from immunocompromised individuals resulted in this sex distribution being less apparent for immunocompromised individuals compared with that seen in the general population (18, 41, 42). Amongst immunocompromised individuals, males accounted for 52.6% of reported events of myocarditis and pericarditis submitted to all three datasets combined, thus a slight decrease in male predominance for this effect. Overall, 59.7% cases of myocarditis and pericarditis reported from immunocompromised individuals were under the age of 64 years. Furthermore, many of the reported events submitted to EudraVigilance and VAERS amongst immunocompromised subgroups met the criteria for a serious case (77.6% of 49 reports submitted to EudraVigilance, and 68.4% of 57 reports to VAERS). Meanwhile, 76.8% of myocarditis or pericarditis cases reported after COVID-19 mRNA vaccines to EudraVigilance from the population as a whole met the criteria for a serious case, while 63.4% of all cases reported to VAERS were serious in nature. For the VAERS population, time-to-onset was consistent with previous suggestions that these events occur within 14 days of vaccination, with approximately 70% of events reported to occur within 14 days of receiving a COVID-19 mRNA vaccine (40, 44). Consistencies in the frequency of events reported and the seriousness of cases for immunocompromised people within the data sources utilised suggests results may be generalisable to other populations in which mRNA vaccines are used. Interestingly, in comparison to the US data which demonstrated that 50% of events in immunocompromised occurred following the second dose of an mRNA vaccination the UK data demonstrates a less apparent separation between doses one and two (31% vs 39.4%), while 23.9% of events occurred following the third dose.

Spontaneous adverse events reported to the Yellow Card scheme in the UK following a COVID-19 mRNA vaccination resulted in higher reporting of myocarditis and pericarditis in immunocompromised individuals as a proportion of the total myocarditis and pericarditis reports compared to immunocompromised individuals in the EU and US databases (7.1% vs 0.84 and 1.86%, respectively). There are many potential reasons for these differences including different reporting systems that may allow for better collection of comorbidities or concomitant medication. Spontaneously reported data from the population as a whole indicated myocarditis and pericarditis occurred predominately in young males. Results from the UK are consistent with those from the EU and US, where data demonstrated a broadening of these susceptible characteristics in immunocompromised individuals. Up to 15 December 2021, 38.4% of reports of myocarditis and pericarditis following COVID-19 mRNA vaccination were from females in the UK; therefore in the immunocompromised population there had been a 1.37-fold higher frequency of myocarditis and pericarditis reports from immunocompromised females (12). The majority of reports occurred following the second dose (39.4%) in the UK which was consistent with reports from the US (54.4% reported following a second dose), although this interpretation should be taken with caution given that the third dose (booster) programme is still ongoing in the UK, with more data likely to be reported in the coming weeks and months. However, immunocompromised people were prioritised to receive their third (booster) dose when the UK booster vaccination programme started in September 2021, therefore it is expected that the majority of people within this subpopulation would have received their third dose by the datalock point of 01 December 2021 (45, 46).

The use of spontaneous reporting systems has some limitations which are important to recognise, including the possibility that these data are underestimated due to underreporting of events and missing information on comorbidities and concurrent medications within individual case reports (47) (5, 48). Additionally, concomitant medication was not commonly reported; medications for cancer and HIV/AIDS may be used for other conditions and may therefore inaccurately represent the number of immunocompromised patients within the dataset. However, for the EudraVigilance population the use of a proxy measure was the only method for identifying reports from people in these patient subgroups. Furthermore, not all immunosuppressive treatments were examined in this analysis, but a comprehensive list of immunosuppressants should be included in future studies. It will therefore become increasingly important to monitor the occurrence and clinical course of myocarditis and pericarditis amongst immunocompromised patients as booster vaccination programmes progress. While COVID-19 vaccination programmes continue to progress to widespread roll-out of third and subsequent booster doses, more immunocompromised individuals will become exposed to COVID-19 mRNA vaccines and some for the first time, particularly in countries where adenovirus vector COVID-19 vaccines were preferentially given to older members of the population and those at higher clinical risk of severe COVID-19 outcomes (many of whom were immunocompromised) for their first two doses. Current data indicates that three doses of mRNA vaccines are safe, which included immunocompromised individuals in the population cohort (49). Consideration should be given to the risk of myocarditis and pericarditis in vaccine recipients who demonstrate low-or non-response to COVID-19 vaccination where additional vaccine doses are warranted; while immunocompromised patient subgroups have generally been shown to elicit sufficient immune response to COVID-19 vaccination, many of those who do respond poorly may be immunocompromised, including cancer patients or transplant recipients (50). Further studies are required to better understand the occurrence of myocarditis and pericarditis following additional mRNA COVID-19 vaccine doses in immunocompromised individuals and for those who have demonstrated limited response to vaccination.

It should also be noted that undiagnosed COVID-19 infection may be an underlying risk factor for myocarditis or pericarditis rather than the vaccine. In the US, it has been demonstrated that risk of cardiac complications is higher following SARS-CoV-2 infection than mRNA vaccination in all age groups for both males and females (51). In the UK, SARS-CoV-2 infection has been found to increase risk of hospitalisation or death from cardiac complications, including myocarditis and pericarditis, and produce a higher number of excess events due to exposure compared with mRNA vaccination (52). Epidemiological studies are warranted to determine whether a similar pattern is seen amongst immunocompromised individuals. Within the data analysed from spontaneous reporting systems for the immunocompromised group, only one case of pericarditis following Pfizer vaccination was identified as also being infected with COVID-19. As immunocompromised individuals accounted for less than 10% of all reports of myocarditis and pericarditis in response to vaccination across the three regions analysed, detecting cases where COVID-19 infection may also have contributed to the reaction will be a small proportion of this limited number of reports. Thus, levels of detecting in this method of analysis may be unable to identify these potentially rare events in this population. It is worth noting that immunocompromised individuals may also be less likely to become infected if they are shielding or maintaining stricter social distancing measures. This raises the very interesting question of whether vaccination associated myocarditis and pericarditis is a consequence of the immune response elicited to generate a robust response that also occurs following infection. Therefore is vaccination preventing these individuals suffering or reducing the severity of myocarditis and pericarditis if they were exposed to the SARS-CoV-2 virus?

Based on current data, immunocompromised individuals reporting myocarditis or pericarditis differed in gender and age distribution compared to the whole population. As risk factors for myocarditis and pericarditis are less clear in the immunocompromised population, it will be important to continue monitoring the occurrence and clinical course of myocarditis and pericarditis in immunocompromised individuals following COVID-19 mRNA vaccines. Further monitoring of events in immunocompromised patients will be of particular importance if mRNA treatments and vaccines continue to be pursued for these conditions. If mRNA vaccines, and potentially other mRNA-based therapeutics, are linked with adverse events in immunocompromised individuals, this may alter their therapeutic potential, may alter treatment regimes, or may stratify patients into different treatment options. It is worth noting that autoimmune disorders, such as arthritis and systemic lupus, have been linked to pericarditis as well as some immunosuppressive medications, including methotrexate (53). Thus, careful evaluation needs to be undertaken, taking into consideration the baseline characteristics of this population to accurately determine the reporting rates related to mRNA vaccination.

It should be noted that our results are based on the best available, though limited, information. Results should be confirmed by quantitative data from formal pharmacoepidemiological studies, to allow accurate calculation of incident risk of myocarditis and pericarditis amongst immunocompromised patients compared with immune competent individuals. Further ongoing monitoring is needed to formally characterise the clinical course of myocarditis and pericarditis following mRNA vaccination amongst immunocompromised patients. It is important to stress that events of myocarditis and pericarditis following COVID-19 vaccination are very rare and the benefits of COVID-19 vaccination continue to outweigh any perceived risks.

In conclusion, myocarditis and pericarditis are very rare events following COVID-19 mRNA vaccination in immunocompromised individuals, and risk factors such as age and sex are less clear in immunocompromised individuals compared to the total population. Slight differences were observed in reporting frequency of these adverse events between the different immunocompromised populations and vaccines, thus continued monitoring may enhance stratification of patients in the future for the most appropriate vaccine type to use.

## Data Availability

All data produced in the present work are contained in the manuscript

## Declarations Transparency statement

The manuscript’s guarantor affirms that the manuscript is an honest, accurate, and transparent account of the study being reported, and that no important aspects of the study have been omitted.

## Ethics approval

Ethics approval was not required.

## Funding

No external funding was received for the preparation of this manuscript.

## Conflicts of interest

All authors have completed the Unified Competing Interest form (available on request from the corresponding author) and declare: The Drug Safety Research Unit (DSRU) is a registered independent charity (No. 327206) associated with the University of Portsmouth. The DSRU receives donations and grants from pharmaceutical companies; however, the companies have no control over the conduct or publication of its studies. The DSRU has received grants to conduct unconditional studies on the Oxford/AstraZeneca COVID-19 vaccine and is in negotiations to receiving grants for conducting CPRD studies for Pfizer, Moderna, and Janssen COVID-19 vaccines. The DSRU has conducted benefit-risk studies on products for COVID-19, including remdesivir, lopinavir/ritonavir, chloroquine and hydroxychloroquine, and convalescent plasma. Professor Shakir is the principal investigator for an active surveillance study for the Oxford/AstraZeneca vaccine, but this assessment is unrelated to this study. Professor Shakir has been a member of Data Safety Monitoring Boards for Ipsen, Biogen, and Diurnal. None of these companies have any involvement with COVID-19 vaccines. Professor Shakir was invited by AstraZeneca to advise on the events of thrombosis with thrombocytopenia with the COVID-19 vaccine and to be a member of an advisory committee on a safety study of the Oxford/AstraZeneca vaccine in Europe. Samantha Lane and Alison Yeomans have no conflicts of interest with regard to this study.

## Authors’ contributions

SL and AY were responsible for data acquisition, analyses, and interpretation. All authors were responsible for study conception, drafting and reviewing the manuscript, and approval of the final version for publication.

## Data sharing

No additional data are available.

## Notes

### Author Declarations

Systematic searches of spontaneous reporting outputs of the European Union/European Economic Area (EU/EEA; EudraVigilance) and the United States (US; Vaccine Adverse Event Reporting System [VAERS] via CDC Wonder tool) were used in this study.

### Summary of Updates

Updated version following peer review

