## Supplementary file for "Systematic review of spontaneous reports of myocarditis and pericarditis in transplant recipients and immunocompromised patients following COVID-19 mRNA vaccination"

Supplementary File to accompany the manuscript entitled "*Systematic review of spontaneous reports of myocarditis and pericarditis in transplant recipients and immunocompromised patients following COVID-19 mRNA vaccination*" by Lane et al.

**Search Strategies used:**

**EudraVigilance:**

Searches were conducted within the EudraVigilance online system (https://www.adrreports.eu/). Searches were conducted for each mRNA COVID-19 vaccine individually to provide a line listing of reported myocarditis and pericarditis events for the overall population.

Symptom terms: "Myocarditis" and "Pericarditis"

Restrictions: European Economic Area geographic origin only

Returned records were then filtered by reported concomitant medications to use as a proxy for immunosuppression, these search terms were identical to those used in the VAERS medications at time of vaccination, please see below. Where both myocarditis and pericarditis were reported by the same case, this was counted as "Myopericarditis."

**MHRA Yellow Card scheme:**

Searches of MHRA Yellow Card records were conducted by MHRA and provided to the research team for further analysis.

Reaction terms (MedDRA PT): Acute endocarditis, Atypical mycobacterium pericarditis, Autoimmune myocarditis, Autoimmune pericarditis, Bacterial pericarditis, Camptodactyly-arthropathy-coxa vara-pericarditis syndrome, Carditis, Coxsackie carditis, Coxsackie endocarditis, Coxsackie myocarditis, Coxsackie pericarditis, Cytomegalovirus myocarditis, Cytomegalovirus pericarditis, Endocarditis, Endocarditis Q fever, Endocarditis bacterial, Endocarditis candida, Endocarditis enterococcal, Endocarditis fibroplastica, Endocarditis gonococcal, Endocarditis haemophilus, Endocarditis helminthic, Endocarditis histoplasma, Endocarditis meningococcal, Endocarditis noninfective, Endocarditis prophylaxis, Endocarditis pseudomonal, Endocarditis rheumatic, Endocarditis staphylococcal, Endocarditis syphilitic, Endocarditis viral, Enterovirus myocarditis, Eosinophilic myocarditis, Fungal endocarditis, Giant cell myocarditis, Hypersensitivity myocarditis, Immune-mediated myocarditis, Lupus endocarditis, Lupus myocarditis, Lyme carditis, Malarial myocarditis, Meningococcal carditis, Myocarditis, Myocarditis bacterial, Myocarditis helminthic, Myocarditis infectious, Myocarditis meningococcal, Myocarditis mycotic, Myocarditis post infection, Myocarditis septic, Myocarditis syphilitic, Myocarditis toxoplasmal, Pericarditis, Pericarditis adhesive, Pericarditis amoebic, Pericarditis constrictive, Pericarditis fungal, Pericarditis gonococcal, Pericarditis helminthic, Pericarditis histoplasma, Pericarditis infective, Pericarditis lupus, Pericarditis malignant, Pericarditis meningococcal, Pericarditis mycoplasmal, Pericarditis rheumatic, Pericarditis syphilitic, Pericarditis tuberculous, Pericarditis uraemic, Pleuropericarditis, Purulent pericarditis, Radiation myocarditis, Radiation pericarditis, Streptococcal endocarditis, Subacute endocarditis, Syphilitic endocarditis of heart valve, Viral myocarditis, Viral pericarditis

Medical History and indication of suspect or other drugs: Transplant, cancer, neoplasm, HIV, AIDs, Immunodeficiency, Immune system disorder, lymphoma, steroid therapy, splenectomy, Systemic lupus erythematosus, Rheumatoid arthritis, Inflammatory bowel disease, Crohn's disease, Ulcerative Colitis

Drugs searched: The data was also reviewed to identify co-suspects or other drugs which were Anti-rejection drugs, Monoclonal anti-bodies and steroids

**VAERS:**

Search terms used for all VAERS searches include the following split into the subheadings available on the VAERS cdc wonder site:

Symptom terms: "Myocarditis" and "Pericarditis"

Vaccine Characteristics: "COVID19 (COVID19 VACCINE)", which was limited to vaccine manufacturer: "MODERNA" and "PFIZER\BIONTECH" with "All Doses"

Location, age and gender: "The United State/Territories/Unknown", Age "All ages" and Sex: "All Genders"

We did not restrict on event characteristics or reporting dates – these fields were unrestricted to include "All".

To identify Transplant recipients, we searched the text fields under the heading of History/Allergies using the terms: "transplant"

To identify Methotrexate treated individuals, we searched the text fields under the heading of Medications at time of vaccination using the term: "methotrexate"

To identify HIV patients, we searched the text fields under the heading of Medications at the time of vaccination using the following approved HIV medications: "abacavir, Ziagen, Emtriva, Epivir, Viread, Retrovir, doravirine, Pifeltro, Sustiva, Intelence, Viramune, Edurant, atazanavir, Reyataz, Prezista, Lexiva, Invirase, Aptivus, enfuvirtide, Fuzeon, maraviroc, Selzentry, cabotegravir, Vocabria, Tivicay, Isentress, fostemsavir, Rukobia, ibalizumab-uiyk, Trogarzo, cobicistat, Tybost, abacavir and lamivudine, Epzicom, Triumeq, Trizivir, Evotaz, Biktarvy, Cabenuva, Prezcobix, Symtuza, Dovato, Juluca, Delstrigo, Atripla, Symfi, Symfi Lo, Genvoya, Stribild, Odefsey, Complera, Descovy, Truvada, Cimduo, Combivir, Kaletra"

To identify Cancer patients, we searched the text fields under the heading of Medications at the time of vaccination using the following approved Cancer medications:

Abecma (Idecabtagene Vicleucel)

Abemaciclib

Abiraterone Acetate

Abraxane (Paclitaxel Albumin-stabilized Nanoparticle Formulation)

ABVD

ABVE

ABVE-PC

AC

Acalabrutinib

AC-T

Actemra (Tocilizumab)

Adcetris (Brentuximab Vedotin)

ADE

Ado-Trastuzumab Emtansine

Adriamycin (Doxorubicin Hydrochloride)

Afatinib Dimaleate

Afinitor (Everolimus)

Akynzeo (Netupitant and Palonosetron Hydrochloride)

Aldara (Imiquimod)

Aldesleukin

Alecensa (Alectinib)

Alectinib

Alemtuzumab

Alimta (Pemetrexed Disodium)

Aliqopa (Copanlisib Hydrochloride)

Alkeran for Injection (Melphalan Hydrochloride)

Alkeran Tablets (Melphalan)

Aloxi (Palonosetron Hydrochloride)

Alpelisib

Alunbrig (Brigatinib)

Ameluz (Aminolevulinic Acid Hydrochloride)

Amifostine

Aminolevulinic Acid Hydrochloride

Amivantamab-vmjw

Anastrozole

Apalutamide

Aprepitant

Aranesp (Darbepoetin Alfa)

Aredia (Pamidronate Disodium)

Arimidex (Anastrozole)

Aromasin (Exemestane)

Arranon (Nelarabine)

Arsenic Trioxide

Arzerra (Ofatumumab)

Asparaginase Erwinia Chrysanthemi

Asparaginase Erwinia Chrysanthemi (Recombinant)-rywn

Asparlas (Calaspargase Pegol-mknl)

Atezolizumab

Avapritinib

Avastin (Bevacizumab)

Avelumab

Axicabtagene Ciloleucel

Axitinib

Ayvakit (Avapritinib)

Azacitidine

Azedra (Iobenguane I 131)

Balversa (Erdafitinib)

Bavencio (Avelumab)

BEACOPP

Belantamab Mafodotin-blmf

Beleodaq (Belinostat)

Belinostat

Belzutifan

Bendamustine Hydrochloride

Bendeka (Bendamustine Hydrochloride)

BEP

Besponsa (Inotuzumab Ozogamicin)

Bevacizumab

Bexarotene

Bicalutamide

BiCNU (Carmustine)

Binimetinib

Blenrep (Belantamab Mafodotin-blmf)

Bleomycin Sulfate

Blinatumomab

Blincyto (Blinatumomab)

Bortezomib

Bosulif (Bosutinib)

Bosutinib

Braftovi (Encorafenib)

Brentuximab Vedotin

Brexucabtagene Autoleucel

Breyanzi (Lisocabtagene Maraleucel)

Brigatinib

Brukinsa (Zanubrutinib)

BuMel

Busulfan

Busulfex (Busulfan)

Cabazitaxel

Cablivi (Caplacizumab-yhdp)

Cabometyx (Cabozantinib-S-Malate)

Cabozantinib-S-Malate

CAF

Calaspargase Pegol-mknl

Calquence (Acalabrutinib)

Campath (Alemtuzumab)

Camptosar (Irinotecan Hydrochloride)

Capecitabine

Caplacizumab-yhdp

Capmatinib Hydrochloride

CAPOX

Carac (Fluorouracil--Topical)

Carboplatin

CARBOPLATIN-TAXOL

Carfilzomib

Carmustine

Carmustine Implant

Casodex (Bicalutamide)

CEM

Cemiplimab-rwlc

Ceritinib

Cerubidine (Daunorubicin Hydrochloride)

Cervarix (Recombinant HPV Bivalent Vaccine)

Cetuximab

CEV

Chlorambucil

CHLORAMBUCIL-PREDNISONE

CHOP

Cisplatin

Cladribine

Clofarabine

Clolar (Clofarabine)

CMF

Cobimetinib Fumarate

Cometriq (Cabozantinib-S-Malate)

Copanlisib Hydrochloride

COPDAC

Copiktra (Duvelisib)

COPP

COPP-ABV

Cosmegen (Dactinomycin)

Cotellic (Cobimetinib Fumarate)

Crizotinib

CVP

Cyclophosphamide

Cyramza (Ramucirumab)

Cytarabine

Dabrafenib Mesylate

Dacarbazine

Dacogen (Decitabine)

Dacomitinib

Dactinomycin

Danyelza (Naxitamab-gqgk)

Daratumumab

Daratumumab and Hyaluronidase-fihj

Darbepoetin Alfa

Darolutamide

Darzalex (Daratumumab)

Darzalex Faspro (Daratumumab and Hyaluronidase-fihj)

Dasatinib

Daunorubicin Hydrochloride

Daunorubicin Hydrochloride and Cytarabine Liposome

Daurismo (Glasdegib Maleate)

Decitabine

Decitabine and Cedazuridine

Defibrotide Sodium

Defitelio (Defibrotide Sodium)

Degarelix

Denileukin Diftitox

Denosumab

Dexamethasone

Dexrazoxane Hydrochloride

Dinutuximab

Docetaxel

Dostarlimab-gxly

Doxil (Doxorubicin Hydrochloride Liposome)

Doxorubicin Hydrochloride

Doxorubicin Hydrochloride Liposome

Durvalumab

Duvelisib

Efudex (Fluorouracil--Topical)

Eligard (Leuprolide Acetate)

Elitek (Rasburicase)

Ellence (Epirubicin Hydrochloride)

Elotuzumab

Eloxatin (Oxaliplatin)

Eltrombopag Olamine

Elzonris (Tagraxofusp-erzs)

Emapalumab-lzsg

Emend (Aprepitant)

Empliciti (Elotuzumab)

Enasidenib Mesylate

Encorafenib

Enfortumab Vedotin-ejfv

Enhertu (Fam-Trastuzumab Deruxtecan-nxki)

Entrectinib

Enzalutamide

Epirubicin Hydrochloride

EPOCH

Epoetin Alfa

Epogen (Epoetin Alfa)

Erbitux (Cetuximab)

Erdafitinib

Eribulin Mesylate

Erivedge (Vismodegib)

Erleada (Apalutamide)

Erlotinib Hydrochloride

Erwinaze (Asparaginase Erwinia chrysanthemi)

Ethyol (Amifostine)

Etopophos (Etoposide Phosphate)

Etoposide

Etoposide Phosphate

Everolimus

Evista (Raloxifene Hydrochloride)

Evomela (Melphalan Hydrochloride)

Exemestane

Exkivity (Mobocertinib Succinate)

5-FU (Fluorouracil Injection)

5-FU (Fluorouracil--Topical)

Fam-Trastuzumab Deruxtecan-nxki

Fareston (Toremifene)

Farydak (Panobinostat Lactate)

Faslodex (Fulvestrant)

FEC

Fedratinib Hydrochloride

Femara (Letrozole)

Filgrastim

Firmagon (Degarelix)

Fludarabine Phosphate

Fluoroplex (Fluorouracil--Topical)

Fluorouracil Injection

Fluorouracil--Topical

Flutamide

FOLFIRI

FOLFIRI-BEVACIZUMAB

FOLFIRI-CETUXIMAB

FOLFIRINOX

FOLFOX

Folotyn (Pralatrexate)

Fostamatinib Disodium

Fotivda (Tivozanib Hydrochloride)

Fulphila (Pegfilgrastim)

FU-LV

Fulvestrant

Gamifant (Emapalumab-lzsg)

Gardasil (Recombinant HPV Quadrivalent Vaccine)

Gardasil 9 (Recombinant HPV Nonavalent Vaccine)

Gavreto (Pralsetinib)

Gazyva (Obinutuzumab)

Gefitinib

Gemcitabine Hydrochloride

GEMCITABINE-CISPLATIN

GEMCITABINE-OXALIPLATIN

Gemtuzumab Ozogamicin

Gemzar (Gemcitabine Hydrochloride)

Gilotrif (Afatinib Dimaleate)

Gilteritinib Fumarate

Glasdegib Maleate

Gleevec (Imatinib Mesylate)

Gliadel Wafer (Carmustine Implant)

Glucarpidase

Goserelin Acetate

Granisetron

Granisetron Hydrochloride

Granix (Filgrastim)

Halaven (Eribulin Mesylate)

Hemangeol (Propranolol Hydrochloride)

Herceptin Hylecta (Trastuzumab and Hyaluronidase-oysk)

Herceptin (Trastuzumab)

HPV Bivalent Vaccine, Recombinant

HPV Nonavalent Vaccine, Recombinant

HPV Quadrivalent Vaccine, Recombinant

Hycamtin (Topotecan Hydrochloride)

Hydrea (Hydroxyurea)

Hydroxyurea

Hyper-CVAD

Ibrance (Palbociclib)

Ibritumomab Tiuxetan

Ibrutinib

ICE

Iclusig (Ponatinib Hydrochloride)

Idamycin PFS (Idarubicin Hydrochloride)

Idarubicin Hydrochloride

Idecabtagene Vicleucel

Idelalisib

Idhifa (Enasidenib Mesylate)

Ifex (Ifosfamide)

Ifosfamide

IL-2 (Aldesleukin)

Imatinib Mesylate

Imbruvica (Ibrutinib)

Imfinzi (Durvalumab)

Imiquimod

Imlygic (Talimogene Laherparepvec)

Infigratinib Phosphate

Infugem (Gemcitabine Hydrochloride)

Inlyta (Axitinib)

Inotuzumab Ozogamicin

Inqovi (Decitabine and Cedazuridine)

Inrebic (Fedratinib Hydrochloride)

Interferon Alfa-2b, Recombinant

Interleukin-2 (Aldesleukin)

Intron A (Recombinant Interferon Alfa-2b)

Iobenguane I 131

Ipilimumab

Iressa (Gefitinib)

Irinotecan Hydrochloride

Irinotecan Hydrochloride Liposome

Isatuximab-irfc

Istodax (Romidepsin)

Ivosidenib

Ixabepilone

Ixazomib Citrate

Ixempra (Ixabepilone)

Jakafi (Ruxolitinib Phosphate)

JEB

Jelmyto (Mitomycin)

Jemperli (Dostarlimab-gxly)

Jevtana (Cabazitaxel)

Kadcyla (Ado-Trastuzumab Emtansine)

Kepivance (Palifermin)

Keytruda (Pembrolizumab)

Kisqali (Ribociclib)

Koselugo (Selumetinib Sulfate)

Kymriah (Tisagenlecleucel)

Kyprolis (Carfilzomib)

Lanreotide Acetate

Lapatinib Ditosylate

Larotrectinib Sulfate

Lenalidomide

Lenvatinib Mesylate

Lenvima (Lenvatinib Mesylate)

Letrozole

Leucovorin Calcium

Leukeran (Chlorambucil)

Leuprolide Acetate

Levulan Kerastik (Aminolevulinic Acid Hydrochloride)

Libtayo (Cemiplimab-rwlc)

Lisocabtagene Maraleucel

Lomustine

Loncastuximab Tesirine-lpyl

Lonsurf (Trifluridine and Tipiracil Hydrochloride)

Lorbrena (Lorlatinib)

Lorlatinib

Lumakras (Sotorasib)

Lumoxiti (Moxetumomab Pasudotox-tdfk)

Lupron Depot (Leuprolide Acetate)

Lurbinectedin

Luspatercept-aamt

Lutathera (Lutetium Lu 177-Dotatate)

Lutetium (Lu 177-Dotatate)

Lynparza (Olaparib)

Margenza (Margetuximab-cmkb)

Margetuximab-cmkb

Marqibo (Vincristine Sulfate Liposome)

Matulane (Procarbazine Hydrochloride)

Mechlorethamine Hydrochloride

Megestrol Acetate

Mekinist (Trametinib Dimethyl Sulfoxide)

Mektovi (Binimetinib)

Melphalan

Melphalan Hydrochloride

Mercaptopurine

Mesna

Mesnex (Mesna)

Methotrexate Sodium

Methylnaltrexone Bromide

Midostaurin

Mitomycin

Mitoxantrone Hydrochloride

Mobocertinib Succinate

Mogamulizumab-kpkc

Monjuvi (Tafasitamab-cxix)

Moxetumomab Pasudotox-tdfk

Mozobil (Plerixafor)

MVAC

Mvasi (Bevacizumab)

Myleran (Busulfan)

Mylotarg (Gemtuzumab Ozogamicin)

Nanoparticle Paclitaxel (Paclitaxel Albumin-stabilized Nanoparticle Formulation)

Naxitamab-gqgk

Necitumumab

Nelarabine

Neratinib Maleate

Nerlynx (Neratinib Maleate)

Netupitant and Palonosetron Hydrochloride

Neulasta (Pegfilgrastim)

Neupogen (Filgrastim)

Nexavar (Sorafenib Tosylate)

Nilandron (Nilutamide)

Nilotinib

Nilutamide

Ninlaro (Ixazomib Citrate)

Niraparib Tosylate Monohydrate

Nivestym (Filgrastim)

Nivolumab

Nplate (Romiplostim)

Nubeqa (Darolutamide)

Nyvepria (Pegfilgrastim)

Obinutuzumab

Odomzo (Sonidegib)

OEPA

Ofatumumab

OFF

Olaparib

Omacetaxine Mepesuccinate

Oncaspar (Pegaspargase)

Ondansetron Hydrochloride

Onivyde (Irinotecan Hydrochloride Liposome)

Ontak (Denileukin Diftitox)

Onureg (Azacitidine)

Opdivo (Nivolumab)

OPPA

Orgovyx (Relugolix)

Osimertinib Mesylate

Oxaliplatin

Paclitaxel

Paclitaxel Albumin-stabilized Nanoparticle Formulation

PAD

Padcev (Enfortumab Vedotin-ejfv)

Palbociclib

Palifermin

Palonosetron Hydrochloride

Palonosetron Hydrochloride and Netupitant

Pamidronate Disodium

Panitumumab

Panobinostat Lactate

Paraplatin (Carboplatin)

Pazopanib Hydrochloride

PCV

PEB

Pegaspargase

Pegfilgrastim

Peginterferon Alfa-2b

PEG-Intron (Peginterferon Alfa-2b)

Pemazyre (Pemigatinib)

Pembrolizumab

Pemetrexed Disodium

Pemigatinib

Perjeta (Pertuzumab)

Pertuzumab

Pertuzumab, Trastuzumab, and Hyaluronidase-zzxf

Pexidartinib Hydrochloride

Phesgo (Pertuzumab, Trastuzumab, and Hyaluronidase-zzxf)

Piqray (Alpelisib)

Plerixafor

Polatuzumab Vedotin-piiq

Polivy (Polatuzumab Vedotin-piiq)

Pomalidomide

Pomalyst (Pomalidomide)

Ponatinib Hydrochloride

Portrazza (Necitumumab)

Poteligeo (Mogamulizumab-kpkc)

Pralatrexate

Pralsetinib

Prednisone

Procarbazine Hydrochloride

Procrit (Epoetin Alfa)

Proleukin (Aldesleukin)

Prolia (Denosumab)

Promacta (Eltrombopag Olamine)

Propranolol Hydrochloride

Provenge (Sipuleucel-T)

Purinethol (Mercaptopurine)

Purixan (Mercaptopurine)

Qinlock (Ripretinib)

Radium 223 Dichloride

Raloxifene Hydrochloride

Ramucirumab

Rasburicase

Ravulizumab-cwvz

Reblozyl (Luspatercept-aamt)

R-CHOP

R-CVP

Recombinant Human Papillomavirus (HPV) Bivalent Vaccine

Recombinant Human Papillomavirus (HPV) Nonavalent Vaccine

Recombinant Human Papillomavirus (HPV) Quadrivalent Vaccine

Recombinant Interferon Alfa-2b

Regorafenib

Relistor (Methylnaltrexone Bromide)

Relugolix

R-EPOCH

Retacrit (Epoetin Alfa)

Retevmo (Selpercatinib)

Revlimid (Lenalidomide)

Ribociclib

R-ICE

Ripretinib

Rituxan (Rituximab)

Rituxan Hycela (Rituximab and Hyaluronidase Human)

Rituximab

Rituximab and Hyaluronidase Human

Rolapitant Hydrochloride

Romidepsin

Romiplostim

Rozlytrek (Entrectinib)

Rubidomycin (Daunorubicin Hydrochloride)

Rubraca (Rucaparib Camsylate)

Rucaparib Camsylate

Ruxolitinib Phosphate

Rybrevant (Amivantamab-vmjw)

Rydapt (Midostaurin)

Rylaze (Asparaginase Erwinia Chrysanthemi [Recombinant]-rywn)

Sacituzumab Govitecan-hziy

Sancuso (Granisetron)

Sarclisa (Isatuximab-irfc)

Sclerosol Intrapleural Aerosol (Talc)

Selinexor

Selpercatinib

Selumetinib Sulfate

Siltuximab

Sipuleucel-T

Soltamox (Tamoxifen Citrate)

Somatuline Depot (Lanreotide Acetate)

Sonidegib

Sorafenib Tosylate

Sotorasib

Sprycel (Dasatinib)

STANFORD V

Sterile Talc Powder (Talc)

Steritalc (Talc)

Stivarga (Regorafenib)

Sunitinib Malate

Sustol (Granisetron)

Sutent (Sunitinib Malate)

Sylatron (Peginterferon Alfa-2b)

Sylvant (Siltuximab)

Synribo (Omacetaxine Mepesuccinate)

Tabloid (Thioguanine)

Tabrecta (Capmatinib Hydrochloride)

TAC

Tafasitamab-cxix

Tafinlar (Dabrafenib Mesylate)

Tagraxofusp-erzs

Tagrisso (Osimertinib Mesylate)

Talazoparib Tosylate

Talc

Talimogene Laherparepvec

Talzenna (Talazoparib Tosylate)

Tamoxifen Citrate

Tarceva (Erlotinib Hydrochloride)

Targretin (Bexarotene)

Tasigna (Nilotinib)

Tavalisse (Fostamatinib Disodium)

Taxotere (Docetaxel)

Tazemetostat Hydrobromide

Tazverik (Tazemetostat Hydrobromide)

Tecartus (Brexucabtagene Autoleucel)

Tecentriq (Atezolizumab)

Temodar (Temozolomide)

Temozolomide

Temsirolimus

Tepadina (Thiotepa)

Tepmetko (Tepotinib Hydrochloride)

Tepotinib Hydrochloride

Thalidomide

Thalomid (Thalidomide)

Thioguanine

Thiotepa

Tibsovo (Ivosidenib)

Tisagenlecleucel

Tivozanib Hydrochloride

Tocilizumab

Tolak (Fluorouracil--Topical)

Topotecan Hydrochloride

Toremifene

Torisel (Temsirolimus)

Totect (Dexrazoxane Hydrochloride)

TPF

Trabectedin

Trametinib Dimethyl Sulfoxide

Trastuzumab

Trastuzumab and Hyaluronidase-oysk

Treanda (Bendamustine Hydrochloride)

Trexall (Methotrexate Sodium)

Trifluridine and Tipiracil Hydrochloride

Trisenox (Arsenic Trioxide)

Trodelvy (Sacituzumab Govitecan-hziy)

Truseltiq (Infigratinib Phosphate)

Truxima (Rituximab)

Tucatinib

Tukysa (Tucatinib)

Turalio (Pexidartinib Hydrochloride)

Tykerb (Lapatinib Ditosylate)

Ukoniq (Umbralisib Tosylate)

Ultomiris (Ravulizumab-cwvz)

Umbralisib Tosylate

Undencyca (Pegfilgrastim)

Unituxin (Dinutuximab)

Uridine Triacetate

VAC

Valrubicin

Valstar (Valrubicin)

Vandetanib

VAMP

Varubi (Rolapitant Hydrochloride)

Vectibix (Panitumumab)

VeIP

Velcade (Bortezomib)

Vemurafenib

Venclexta (Venetoclax)

Venetoclax

Verzenio (Abemaciclib)

Vidaza (Azacitidine)

Vinblastine Sulfate

Vincristine Sulfate

Vincristine Sulfate Liposome

Vinorelbine Tartrate

VIP

Vismodegib

Vistogard (Uridine Triacetate)

Vitrakvi (Larotrectinib Sulfate)

Vizimpro (Dacomitinib)

Voraxaze (Glucarpidase)

Vorinostat

Votrient (Pazopanib Hydrochloride)

Vyxeos (Daunorubicin Hydrochloride and Cytarabine Liposome)

Welireg (Belzutifan)

Xalkori (Crizotinib)

Xatmep (Methotrexate Sodium)

Xeloda (Capecitabine)

XELIRI

XELOX

Xgeva (Denosumab)

Xofigo (Radium 223 Dichloride)

Xospata (Gilteritinib Fumarate)

Xpovio (Selinexor)

Xtandi (Enzalutamide)

Yervoy (Ipilimumab)

Yescarta (Axicabtagene Ciloleucel)

Yondelis (Trabectedin)

Yonsa (Abiraterone Acetate)

Zaltrap (Ziv-Aflibercept)

Zanubrutinib

Zarxio (Filgrastim)

Zejula (Niraparib Tosylate Monohydrate)

Zelboraf (Vemurafenib)

Zepzelca (Lurbinectedin)

Zevalin (Ibritumomab Tiuxetan)

Ziextenzo (Pegfilgrastim)

Zinecard (Dexrazoxane Hydrochloride)

Zirabev (Bevcizumab)

Ziv-Aflibercept

Zofran (Ondansetron Hydrochloride)

Zoladex (Goserelin Acetate)

Zoledronic Acid

Zolinza (Vorinostat)

Zometa (Zoledronic Acid)

Zyclara (Imiquimod)

Zydelig (Idelalisib)

Zykadia (Ceritinib)

Zynlonta (Loncastuximab Tesirine-lpyl)

Zytiga (Abiraterone Acetate)
